# Patient ABO blood type is a major predictor of a positive DAT following a transfusion reaction

**DOI:** 10.1101/2021.11.01.21265756

**Authors:** Eric Schnieders, Judith Leon, C. Michael Knudson

**Author notes:** Corresponding Author: C. Michael Knudson, Department of Pathology, University of Iowa Hospitals and Clinics, 200 Hawkins Dr., C250 GH.

## Abstract

**Background:** A direct antiglobulin test (DAT) checks for antibody or complement on the surface of RBCs and is often done following a transfusion reaction. While passive anti-A and anti-B antibodies are known to cause positive DATs, the extent this occurs following transfusion is unknown.

**Study Design and methods:** DAT results, ABO type and blood product information was recorded on 1097 transfusion reactions at a large academic hospital over 8 years. The effect of patient blood type, product type and plasma compatibility of blood product transfused on DAT results were determined. Statistical significance was determined using Chi-squared testing.

**Results:** Plasma compatibility of the product was a strong predictor of a positive DAT with plasma compatible transfusions having a 9.4% positive rate while plasma incompatible transfusions were positive 44% of the time (P<0.0001). Patient ABO blood type was a strong predictor of a positive DAT with Type O patients having 6.6% positive rate and non-O patients having a positive rate of 20.6% (P <0.0001). These results were significant for individual blood types as well. Type A, B or AB patients had higher DAT positive rates even when plasma incompatible transfusions were excluded from the analysis (P<0.0001). Platelets were significantly more likely to be associated with a positive DAT when compared to RBC transfusions.

**Conclusions:** These results show plasma compatibility and ABO types are strong predictors of positive DAT results following a transfusion reaction. Anti-A and anti-B antibodies are estimated to account for about 50% of positive DATs in this study.

## INTRODUCTION

The direct antiglobulin test (DAT) is widely used in transfusion medicine^1^. The test is most commonly done with reagents that can detect IgG, complement (C3) or both (polyspecific) on the surface of patients red blood cells (RBCs)^2^. When a transfusion reaction is reported, one of the critical tasks of the blood bank is to assure an immune mediated hemolytic reaction has been excluded and the DAT is commonly used for this purpose. When the DAT is negative shortly after a transfusion reaction, this indicates that an immune (antibody) mediated reaction is unlikely. A positive DAT may indicate that an antibody mediated reaction may have occurred, and further evaluation may be warranted^1-3^.

Many causes for a positive DAT have been identified. These include autoimmune hemolytic anemia, drug-induced hemolytic anemia, hemolytic disease of the newborn, RBC alloantibodies from prior transfusions, hematopoietic stem cell transplantation, and ABO-incompatible organ transplant^2^. Even allogeneic blood donors and healthy control populations have been found to have a positive DAT^4,5^. Drugs such as intravenous immunoglobulin (IVIG) anti-CD38 and anti-CD47 antibodies can cause a positive DAT^6,7, 8^. Because the differential of a positive DAT is so broad, determining the cause of a positive DAT following a transfusion reaction can sometimes be difficult.

As our blood bank policy is to perform DAT on all transfusion reactions, many positive DATs in patients following transfusion reaction were observed that seemed to occur more frequently in patients with A, B or AB blood type. This observation is not surprising to experienced transfusion medicine specialists. It was surprising that a detailed review of the literature identified very few papers addressing the rates of DAT positivity in select patient populations or for select products. One large study involved over 65,049 patients prior to transfusion and found that only 3570 (5.4%) were DAT positive^9^. This study did not look at DAT results after transfusion. Another study of cord blood samples found that the DAT was rarely positive (2.59%) and that most (91%) or these positive results were due to anti-A or anti-B antibodies^10^. Another study published in the 1970s described 47 patients who had a hemolytic transfusion reaction and a positive DAT was very common (42 patients) in this series but this is not surprising in this patient population^11^. Another study looked at allo-antibody formation and DAT results in hemodialysis patients receiving RBC transfusions and found that the DAT was positive in about 15% of patients but found only one patient with anti-A,B in the eluate^12^. Another study looked at DAT results in patients suspected of having delayed hemolytic or delayed serological transfusions reactions but this study focused only on these patients which are likely to have a positive DAT and did not examine the role of anti-A or anti-B antibodies in these patients^13^. No studies were found that looked at DAT positive rates in unselected patients following any transfusion reaction.

Our long standing policy to perform DATs on all transfusion reactions provides an opportunity to tabulate those results and determine the major predictors of a positive DAT following a transfusion reaction. In this study, we hypothesized that passively transfused anti-A, and anti-B found in the plasma of transfused products would be a common cause of positive DATs in patients being evaluated following a transfusion reaction. DAT results from over 1000 transfusion reaction workups were tabulated to test this hypothesis and to determine how frequent positive DAT results are in this patient population.

## MATERIALS AND METHODS

### Study Overview

This retrospective study was approved by the institutional review board (IRB#202001338) at our large academic medical center, which encompasses a level 1 trauma center, solid organ transplantation, hematopoietic stem cell transplantation, and a pediatric hospital with a neonatal intensive care unit. Blood bank records of all transfusion reactions reported over an eight year period from January 2012 to Dec 2019 were reviewed. We recorded information about the patient, the type of transfusion and the transfusion reaction workup including DAT results for each of these reactions. Electronic medical records (EMR; EPIC Systems Corporation) was reviewed if necessary when information was not found in blood bank records.

### Transfusion reactions

The dataset utilized in this study included all suspected transfusion reactions reported to the blood bank during the study period. Per protocol at our institution, when a suspected transfusion reaction is ordered, a patient blood sample is drawn and sent to the blood bank to repeat ABO-Rh blood typing and perform a direct antiglobulin test (DAT). The transfusion service is AABB accredited and participates in College of American Pathology (CAP) surveys that includes blinded DAT samples. The DAT is performed by laboratory personal with a polyspecific (IgG and C3) reagent following standard laboratory procedures. If positive, the DAT is performed with each reagent individually and a DAT is performed on the pre-transfusion sample (type and screen) if a sample is available. A post-transfusion specimen of the patient’s plasma is also examined for evidence of hemolysis.

Suspected transfusion reactions are classified by physicians on the transfusion medicine service. Starting in 2015, the Centers for Disease Control and Prevention (CDC) and National Healthcare Safety Network (NHSN) guidelines for classification of transfusion reactions have been followed. Reactions prior to that time were not reviewed to determine if they met these guidelines. Results for all transfusion reactions were manually entered into a spreadsheet for further analysis.

A total of 1101 transfusion reactions were reported during the study period for 879 unique patients and a post transfusion DAT was performed on 1097 (99.6%) of these reactions. Only the reactions with a DAT performed will be evaluated for this study. The products transfused for these 1097 transfusions reaction workups included packed red blood cells (pRBCs; N=641), platelets (N=376), plasma (N=29), multiple products (N=27), hematopoietic stem cells (N=16), cryoprecipitate (N=5), and granulocytes (N=3). Transfusion reactions reported for these 1097 reactions were as follows: febrile non-hemolytic (FNHTR; N=339), allergic/anaphylactic (N=258), transfusion-associated circulatory overload (TACO; N=78), hypotensive (N=19), transfusion-associated dyspnea (TAD; N=16), acute hemolytic (AHTR; N=13), transfusion-associated acute lung injury (TRALI; N=1), and delayed hemolytic (DHTR; N=1). Other transfusion reactions reported included dimethyl sulfoxide toxicity (N=3), hyperhemolysis (N=2), citrate toxicity (N=1), hyperkalemia (N=1). A total of 366 reported reactions were classified as unknown/unrelated to transfusion.

### Transfusion Compatibility

RBC, platelet, HPC, and granulocyte transfusions were classified as ABO identical (compatible), antigen incompatible (major), plasma incompatible (minor) or both antigen and plasma incompatible (bidirectional). Non-cellular products such as plasma and cryo do not contain significant ABO antigens were all plasma compatible so all of these transfusions were classified as compatible. Transfusion reactions involving multiple products (N=27) were classified based on whether any of the products provided were incompatible using the same definitions as above. Using these definitions, the 1097 transfusion reactions with a post transfusion DAT were classified as compatible (N=851), minor incompatible (N=122), major incompatible (N=98) and bidirectional incompatible (N=26).

### Statistical analysis

Differences in DAT positivity rates were tested for statistical significance using the chi-Square test^14^ as previously described^15^. This test can be applied when the “expected” number of events for each group was at least five^14^. The expected number of events was calculated based on the positive rate for the lowest group being studied. For example, when the effect of blood type is compared, the expected rate of reactions is based on Type O patients (6.7%) since they had the lowest rate among any of the blood types. P-values less than 0.05 were considered statistically significant.

## RESULTS

During this study, 159 positive DATs were recorded following a transfusion reaction giving an overall positive rate of 14.5% (**Table 1**). Patients with blood type O never receive plasma incompatible for ABO as they do not express the A or the B antigen. Thus, if anti-A and anti-B antibodies are a common cause of positive DAT then Type O patients would be expected to have the lowest rate of DAT positivity when compared to patients with other blood types. Only 32 of 480 type O patients (6.7%) were found to have a positive DAT after their transfusion reaction. Type A, B, and AB patients all had a significantly (P<0.0001) higher rate of DAT positivity than the Type O patients (**Table 1**). When type A, B and AB patients are combined, 127 of the 617 DATs performed were positive (20.6%). If this group had the same DAT positive rate as Type O patients then only 41 DAT would have been expected to be positive. This indicates that type A, B or AB patients are roughly 3 times as likely to have a positive DAT following a transfusion reaction than a type O patient. Anti-A and/or anti-B antibodies are a likely cause of the increased DAT positive rate in Type A,B or AB patients. Platelets have much more plasma than RBCs so if passive anti-A and anti-B antibodies account for the increased rates of positive DATs type A,B or AB patients then DAT positive rates might be expected to be even higher with platelet transfusions. Analysis of DAT positivity rates based on patient blood type and product transfused (all products, reactions associated with RBCs or reactions associated with platelets) is shown in **Figure 1**. For the B or AB patients, DAT positive rates were significantly higher following platelet transfusion when compared to RBC transfusions. For type O or A patients the DAT rates did not change substantially based on product transfused. When patients of all blood types are examined, positive DAT were observed in 12.8% of the reactions with RBCs transfused. In contrast, positive DATs were observed in 17% of reactions with platelets transfused. These results were significantly different (P= 0.021).

**Table 1:**
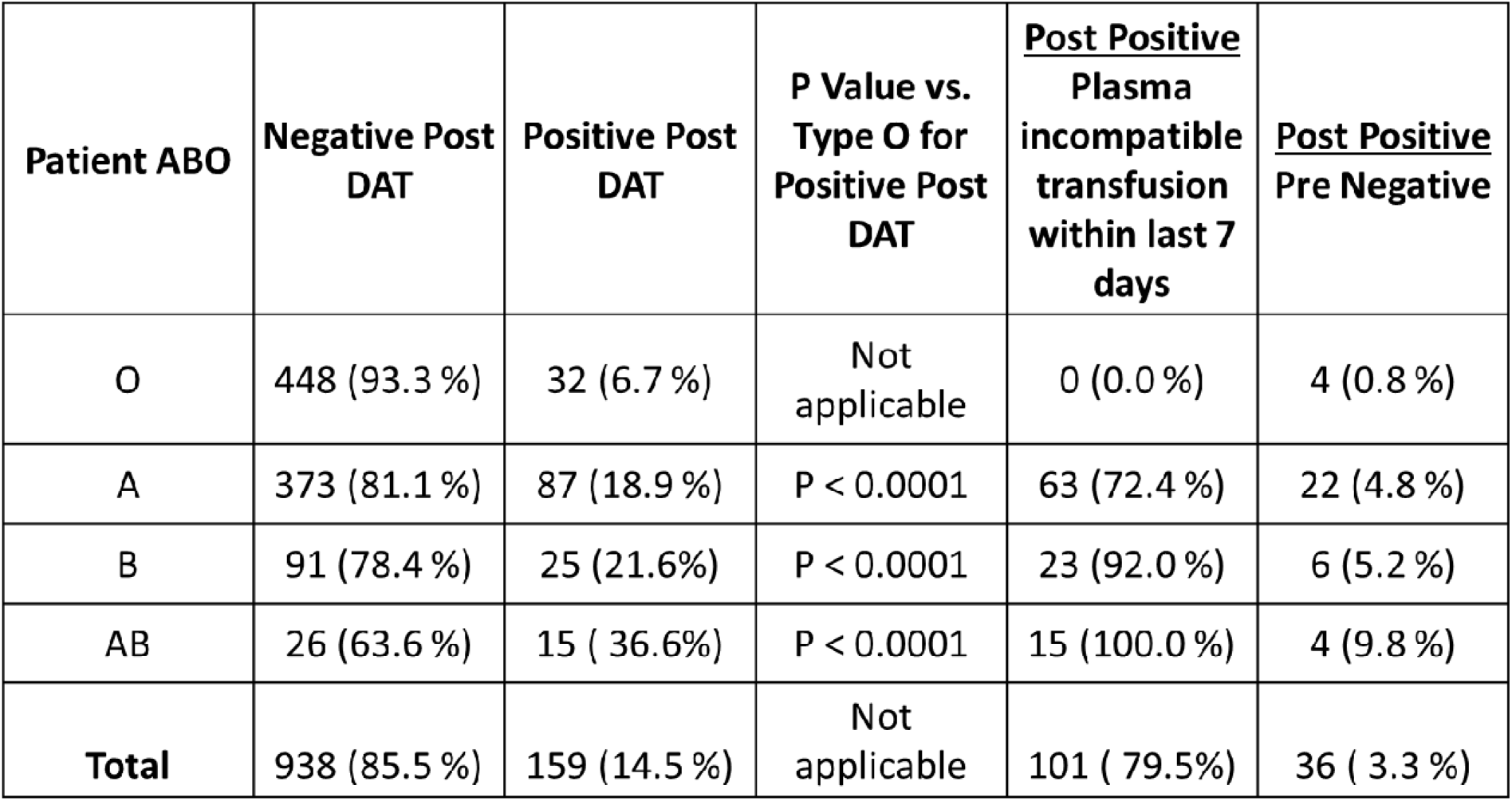
DAT positivity rates for all transfusion reactions by patient ABO type and plasma compatibility.

**Figure 1:**
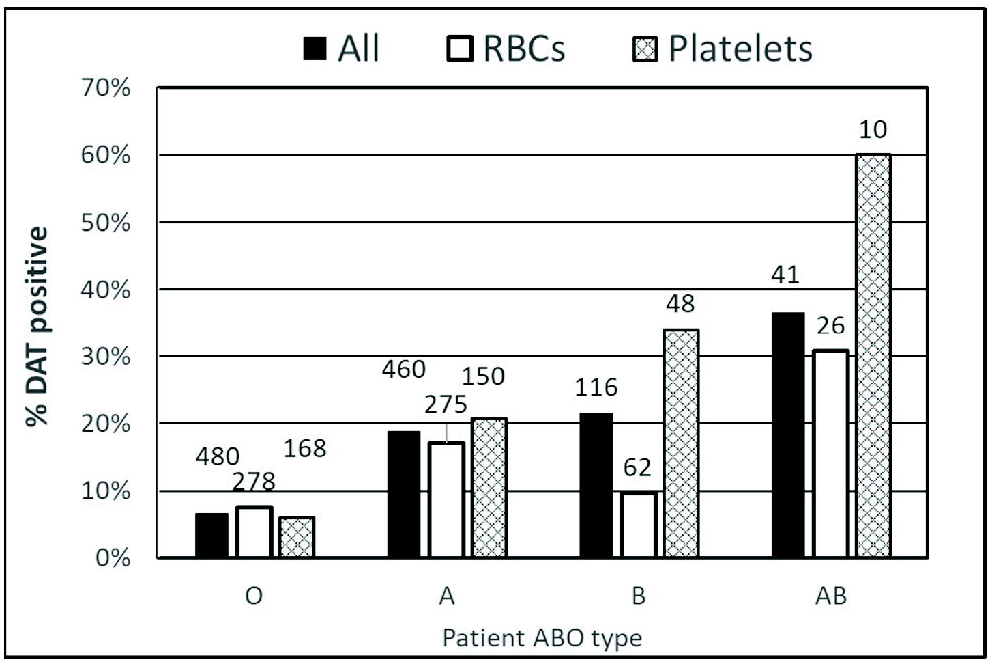
Patient ABO type is a strong predictor of a positive DAT following transfusion reactions with all products, RBCs or Platelets. DAT positivity by patient ABO type for reactions associated with all blood products transfused (N=1097, solid black bar), RBC transfusion RBC (N=641, open bar) or reaction associated with a platelet transfusion (N=376, hatched bars). The number of patients in each group is indicated above each bar. Statistical analysis was performed comparing DAT positive rates in type A, B or AB patients to the type O patients and all results were statistically significant (P<0.01) except for the Type B patients who received RBC products (P=0.54).

Given the significant differences in DAT positivity rates based on patient ABO blood type and on product type, the effect of plasma compatibility of the transfusion on DAT positive rates were determined. Looking at all the transfusion reactions, 951 transfusions were plasma compatible and 9.9% of these were associated with a positive DAT. In contrast, there were 146 transfusions associated with incompatible plasma and 44.5% of these had a positive DAT (**Figure 2A**). Looking at DAT positive rates associated with specific products, both plasma incompatible RBCs (37%) (**Figure 2B**) and plasma incompatible platelets (**Figure 2C**) had much higher DAT positive rates than plasma compatible controls. All these results were statistically different (P<0.001) than what is seen with plasma compatible transfusions. These data strongly suggest that passive anti-A and anti-B antibodies are a common cause of a positive DAT following a transfusion reaction.

**Figure 2:**
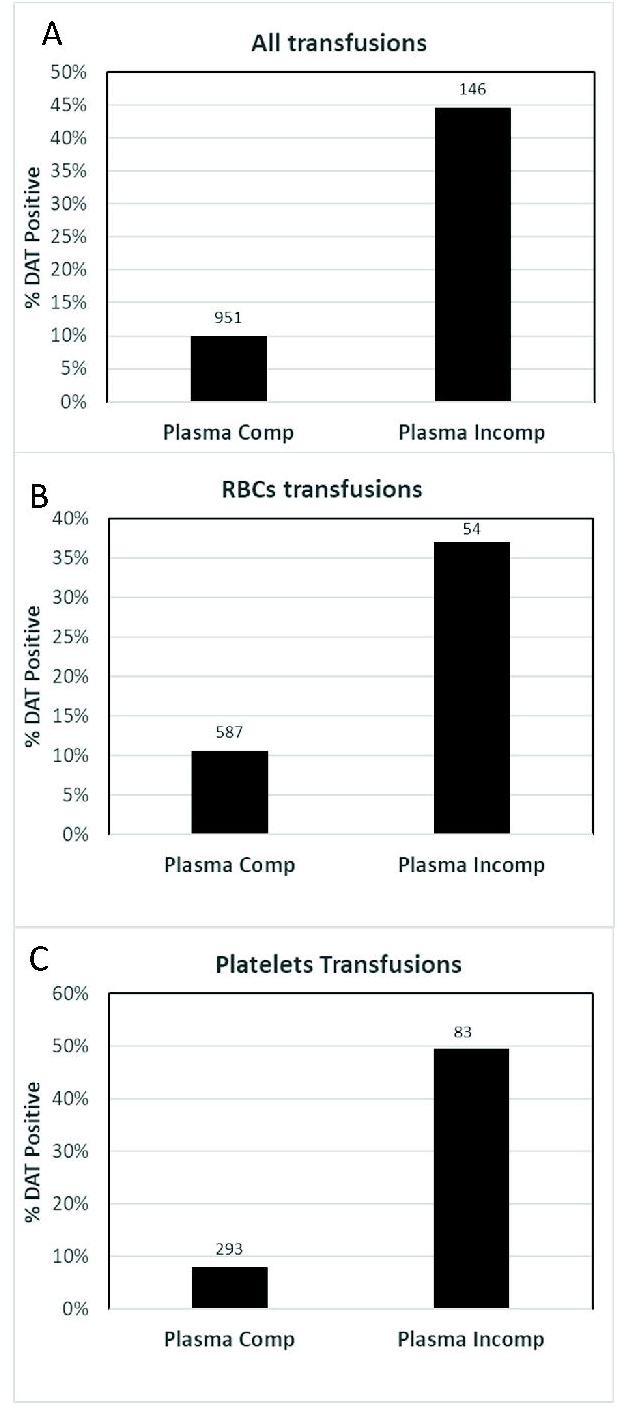
Plasma compatibility is a strong predictor of positive DAT results. **A**) DAT results from all products transfused (N=1097) based on whether the products were plasma compatible or plasma incompatible product is shown. A significantly increased rate of positive DATs was observed when plasma incompatible units were transfused (p<0.0001). **B**) DAT results from patients who were transfused with pRBCs (N=641) based on whether the products were plasma compatible or plasma incompatible product is shown. A significantly increased rate of positive DATs was observed when plasma incompatible units were transfused (p<0.0001). **C**) DAT results from patients who were transfused with platelets (N=376) based on whether the products were plasma compatible or plasma incompatible product is shown. A significantly increased rate of positive DATs was observed when plasma incompatible units were transfused (p<0.0001).

The effect of ABO blood type on DAT positive rates was also examined for plasma compatible transfusions. When plasma incompatible transfusions were excluded, patient blood type still impacted DAT positive rates have an effect as 6.7% of type O patients had positive DATs compared to 13.2% (62 of 471) of type A,B or AB patients. The medical records of these 62 type A,B or AB patients with a positive DAT was examined to determine whether plasma incompatible products were transfused in the 7 days prior to the reaction for the patients with a positive post transfusion DAT. We found just 26 of these A,B or AB patients had not received plasma incompatible transfusions in the previous 7 days (Table 1). If patients receiving incompatible plasma transfusions were excluded from the analysis, 26 or 409 (6%) type A,B or AB patients had a positive DAT following a transfusion reactions. This value matches closely with the rate observed in type O patients. These results suggest that anti-A and anti-B antibodies should be considered as a cause of a positive DAT in type A, B or AB patients even when the implicated unit is plasma compatible with the patient.

## DISCUSSION

The DAT test remains a standard test for the workup of patients following a transfusion reaction. While many hospitals may not routinely perform the DAT test on all transfusions, our long-standing policy to perform this test for all reported reactions allowed us to systematically review the results of DAT testing following any transfusion reaction. We identified multiple factors that predict a positive post-transfusion DAT in reported transfusion reactions. These include patient ABO blood type, plasma compatibility of the product and the product transfused. All these factors may prove useful to both clinicians and transfusion medicine physicians in the work-up of suspected transfusion reactions.

These results demonstrate that type A, B or AB patients have significantly increased rates of positive posttransfusion DATs compared to O patients. Unfortunately, elution studies looking for anti-A and/or anti-B antibodies were not systematically performed for positive DATs during this study period. Nonetheless, the strong relationship between blood type and plasma compatibility of the blood product transfused and DAT positive rates provides strong indirect evidence that many of these are due to passive anti-A and anti-B antibodies. Of note, when type A, B or AB patients receiving plasma incompatible transfusions within the past week were excluded from the analysis, their DAT positive rates were nearly identical to the rate seen in type O patients. These results strongly support our hypothesis that passive transfusion of anti-A and anti-B in plasma incompatible transfusions is a common cause of a positive DAT.

The observation that passive anti-A or anti-B antibodies can cause a positive DAT is certainly not unexpected to an experienced transfusion medicine practitioner. This study examined DAT results on over 1000 transfusion reactions involving more than 800 patients and to our knowledge represents the largest study published on this topic to date. In fact, the second largest study we identified involved just 47 transfusion reactions and focused exclusively on patients who experienced a hemolytic transfusion reaction^11^. This study was performed over 40 years ago and focused exclusively on patients being evaluated for a hemolytic transfusion reaction. While the DAT positive rate was very high in this study (42 of 47), ABO compatibility was not a major risk factor of a hemolytic transfusion reaction at that time as ABO incompatibility only accounted for six of the reactions^11^. To our knowledge, the frequency of positive DATs in patients being evaluated following any transfusion reaction has never been systematically studied.

While this study represents results from just a single center, it represents a compilation of nearly 8 years of results during which time all transfusion reactions are worked up the same regardless of the patient’s symptoms. The results provide strong evidence that incompatible plasma is a common cause of a positive DAT in this setting. In fact, if A,B and AB patients had DAT positive rates that were the same as type O patients in this study (6.7%), 54% fewer positive DATs would have been expected suggesting that more than half of the positive DATs are due to passive anti-A or anti-B antibodies in this study. These results remind us that anti-A and anti-B antibodies are a frequent cause of a positive DAT and that ABO type of the patient and the product should be carefully considered whenever a positive DAT is detected in patient following a transfusion reaction.

## Data Availability

Deidentified data produced in the present study are available upon reasonable request to the authors

## Notes

**Source of support:** Department of Pathology, University of Iowa

### Competing Interest Statement

The authors have declared no competing interest.

### Funding Statement

This study did not receive any funding

### Author Declarations

HawkIRB IRB#202001338

